# Epidemiologic Moderators of the Effectiveness of Routine Screening for LAIs in High-Biosafety Environments

**DOI:** 10.64898/2026.04.05.26350204

**Authors:** Byron Lev Mentis Cohen, William Hanage, Nicolas Menzies, Kevin Croke

**Affiliations:** Harvard Chan School of Public Health

## Abstract

**Justification:** Accidental lab-acquired infections (LAIs) with potential pandemic pathogens (PPPs) in high-biosafety research facilities risk causing a pandemic. Routine testing of lab workers for LAIs coupled with isolation of infected workers could reduce the risk, but the impact of such an intervention may depend on pathogens’ epidemiological characteristics.

**Objective:** This study aims to understand how the epidemiological characteristics of PPPs moderate the efficacy of a routine testing and isolation intervention in preventing larger outbreaks after an LAI.

**Methods:** We employed a discrete-time stochastic network infectious disease model to run 625,000 epidemic simulations encompassing 625 unique combinations of five parameters of interest: test frequency, pathogen transmissibility, the self-isolation rate for symptomatic cases, the percentage of cases that are asymptomatic, and the percentage of infectious time that is spent in the pre-symptomatic state among those who show symptoms. To summarize the Monte Carlo simulations, we paired visual analysis with logistic regression for formal hypothesis testing, with an emphasis on the interaction terms that capture the moderating effect of epidemiological parameters on the impact of test frequency.

**Main Results:** There were four main findings. First, the relative reductions in risk of outbreak that were caused by increased test frequency were inversely correlated with pathogen transmissibility. Second, the effect of test frequency was magnified at higher asymptomatic shares when the symptomatic self-isolation rate was high, but minimally when the self-isolation rate is low. Third, the direction of how the symptomatic self-isolation rate moderated the effect of increased test frequency depended on the asymptomatic share. Fourth, as the pre-symptomatic share of infectious time increased, the effect of test frequency on the probability of an outbreak was strongly magnified largely independent of symptomatic self-isolation rates.

**Conclusions:** Routine testing and isolation could significantly mitigate the risk of catastrophic PPP escapes, with the intervention’s success varying based on pathogen characteristics. High shares of asymptomatic and pre-symptomatic transmission notably increased the relative risk reductions achieved by the intervention. These findings suggest prioritizing testing interventions for pathogens with high asymptomatic and pre-symptomatic transmission and highlight the symptomatic self-isolation rate as a policy intervention target.

## Introduction

In recently published research, Cohen et al found that routine screening and isolation of lab workers in high-biosafety research environments for accidental lab-acquired infections (LAIs) substantially reduced the modeled risk of a catastrophic escape of a potential pandemic pathogen (PPP) similar to wild type SARS-COV-2.^1^ This research paper investigates how the efficacy of such a routine screening intervention might be mediated by the epidemiologic characteristics of the pathogens under study. In particular, this analysis examined how the success of such a surveillance protocol would depend on pathogen transmissibility, the self-isolation rate for symptomatic cases, the percentage of cases that are asymptomatic, and the percentage of infectious time that is spent in with pre-symptomatic infectious disease.

This research is motivated by the need to identify appropriately targeted interventions to mitigate the risk that an accidental lab-acquired infection (LAI) could spark a pandemic.^2–5^ In the forty year period from 1975 to 2016, there were more than 50 documented unintentional human exposures to high-consequence pathogens.^6^ These incidents included exposures to a wide variety of potential high-consequence pathogens including *Variola virus*, various influenza viruses, *SARS-COV-1*, *Ebolavirus*, *Yersinia pestis*, and weaponized *Bacillus anthracis.* ^6^ These and other potential high-consequence pathogens exhibit a wide range of epidemiological characteristics, with major differences in transmissibility, mode of transmission, generation time, presence of asymptomatic or pre-symptomatic transmission, susceptibility to medical countermeasures, likelihood of self-isolation upon developing symptoms, and virulence, among other factors. These epidemiological factors influence the degree to which a pathogen is controllable.^7^ For this reason, it is likely that the efficacy of a program of routine screening of lab workers in high-biosafety research environments for LAIs would vary substantially by pathogen type. Moreover, biosafety interventions in high-biosafety research facilities can in some cases limit the scope of or slow the implementation of research with public health value,^3^ while the resources available for biosafety programs are not unlimited, so it is valuable to gain information allowing for more precise targeting of future routine screening programs to pathogens with the appropriate epidemiological characteristics.

The first epidemiologic characteristic that we investigated as a potential moderator of the intervention’s effectiveness is the pathogen’s transmissibility. We selected this characteristic for analysis because we hypothesized that as pathogen transmissibility rises, the difficulty of pathogen containment rises nonlinearly, meaning that the relative risk reductions of an outbreak that could be achieved with increased test frequency would decline. For example, an intervention generating a 50% reduction in transmission of a pathogen with a basic reproductive number (R_0_) of 1.5 is likely to bring an initial outbreak of that pathogen under control. In contrast, such an intervention alone would not likely end an initial outbreak of a pathogen with a basic reproductive number (R_0_) of 2.5, as a 50% reduction in transmissibility would still leave the effective reproductive number (R_e_) above the crucial threshold of 1.

The second epidemiologic characteristic that we investigated as a potential moderator of the intervention’s effectiveness was the self-isolation rate for symptomatic cases. We selected this parameter for analysis because we hypothesized that since self-isolation and test-induced isolation are competing risks, the effect of test frequency on the probability of an outbreak should be influenced by the self-isolation rate of symptomatic infectees. In a context where the symptomatic self-isolation rate is very high and a large proportion of cases exhibit symptoms, there should be fewer symptomatic cases available to be isolated after testing positive. In addition, we selected this parameter in part because it is a behavioral factor that might be amenable to policy interventions.

The third epidemiologic characteristic that we investigated as a potential moderator of the intervention’s effectiveness was the percentage of cases that are asymptomatic. We selected this parameter for analysis because we hypothesized that since fully asymptomatic people won’t self-isolate due to symptoms, when a larger percentage of cases are asymptomatic there will be more cases available for testing to detect. In effect, having a high percentage of cases that are asymptomatic should reduce the competing risk of self-isolation relative to test-induced isolation. However, the size of the change in the effect of the test intervention resulting from changing the percentage of cases that are asymptomatic would be expected to also depend on the symptomatic self-isolation rate.

The fourth epidemiologic characteristic that we investigated as a potential moderator of the intervention’s effectiveness was the percentage of infectious time that is spent in the pre-symptomatic state, among those who show symptoms. We selected this parameter for analysis because we hypothesized that increasing this percentage would increase the proportion of cases who isolate as the result of a positive test rather than self-isolate after experiencing symptoms, thereby likely augmenting the effect of the testing intervention even if transmissibility is held constant across pre-symptomatic vs symptomatic phases of infection. In effect, having a high percentage of infectious time that is spent in the pre-symptomatic state reduces the competing risk for isolation posed by symptom-induced isolation relative to test-induced isolation. However, the size of this change would also be expected to depend on the symptomatic self-isolation rate.

## Methods

### Modeling Framework and Analytic Scenarios

This analysis was conducted with the discrete-time stochastic network infectious disease model described in Cohen et al (2026), shown in Figure 3.1.^1^ 625 unique combinations of five parameters of interest were generated using a Latin hypercube sample drawn from a plausible range of each parameter being explored. Latin hypercube sampling was selected as a sampling strategy for its desirable coverage properties.^8^ These parameters of interest were test frequency, pathogen transmissibility, the self-isolation rate for symptomatic cases, the percentage of cases that are asymptomatic, and the percentage of infectious time that is spent in the pre-symptomatic state. Routine testing frequency for lab workers was the only variable driving the intensity of the testing regime; all other characteristics of the testing regime were held fixed.

**Figure 3.1:**
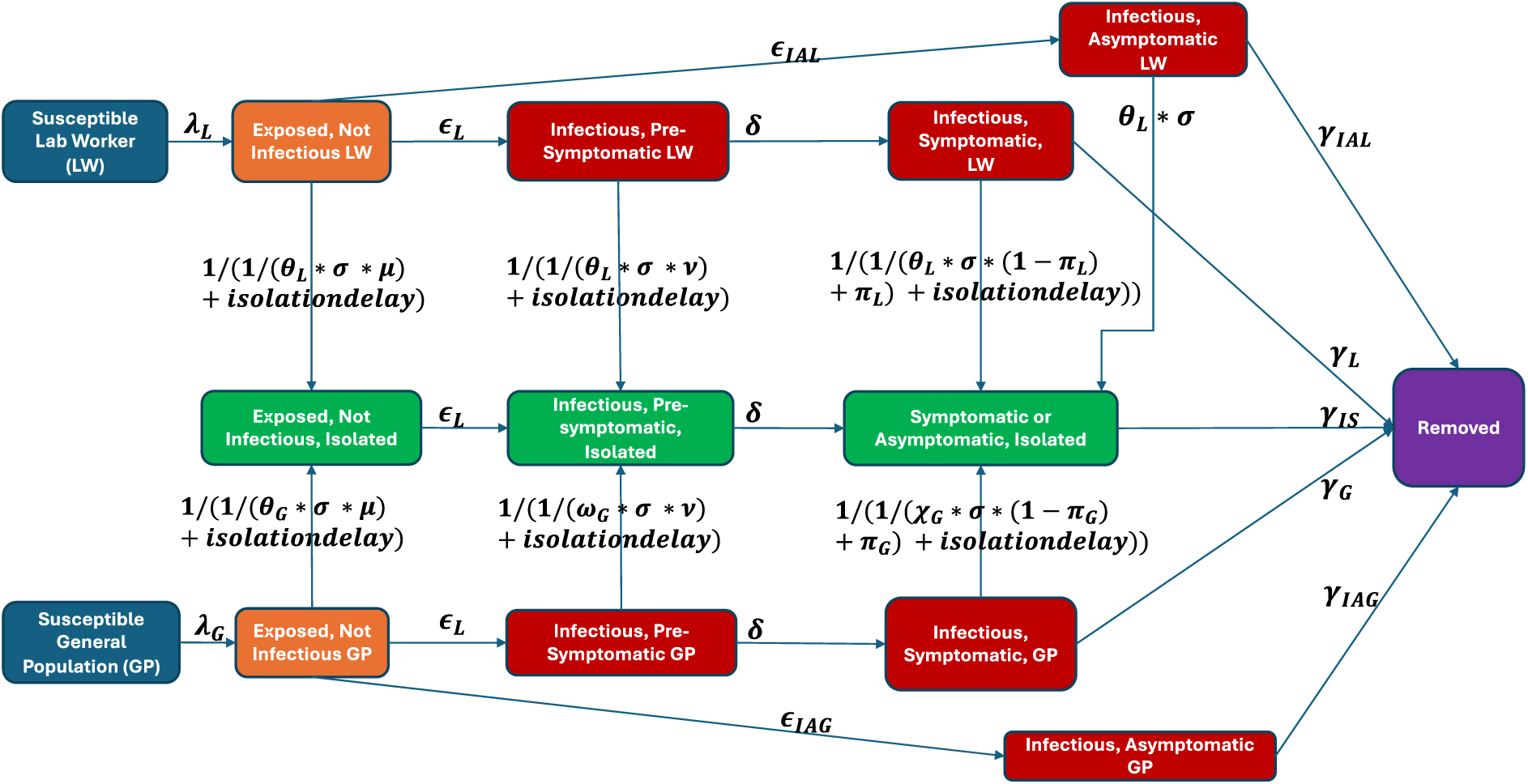
Model Structure.

We conducted 1,000 simulations for each of the 625 unique parameter combinations, for a total of 625,000 simulations. Each simulation was initialized with a single randomly chosen lab worker becoming infected and continued for 100 days. The test frequency ranged from between zero and four tests per week, with peak test sensitivity fixed at 80%, and an average delay from positive test to isolation of one day. To minimize model complexity, tests were assumed to be 100% specific, with no false positives. After the first positive test result, the testing cadence for lab workers increased to daily. In this model, pathogen transmissibility was operationalized by tau, the per-time step probability of transmission across a given pairing of susceptible and infectious individuals who come into contact. To summarize the relationship between tau and R0, we also conducted 5,000 additional simulations with no routine testing that systematically varied only tau while fixing the asymptomatic share at 0.5, the pre-symptomatic share of infectious time at 0.5, and the symptomatic self-isolation rate per day at 0.4, and then calculated mean and median R0 values for each value of tau during weeks one and two of the simulations using a published approach.^9^ The analysis was conducted in R version 4.2.2.^10^

### Statistical and Visual Analysis

We used logistic regression to predict whether or not an outbreak of 50 or more infections would occur and used these statistical models for both hypothesis-testing and graphical interpretation. The logistic regression model took the following form:

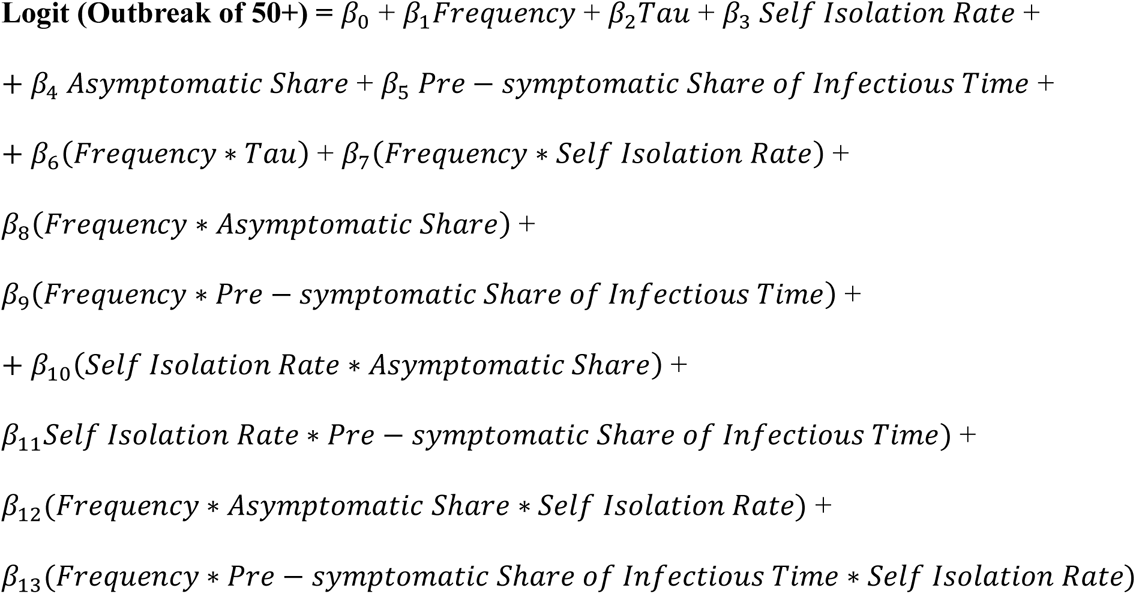

The coefficients of interest were the interaction terms that captured how the effect of test frequency is moderated by epidemiological parameters. However, interpreting the coefficients of a logistic regression model with interaction between continuous terms is difficult, particularly in the presence of three-way interactions.^11^. For this reason, we reported results graphically, to supplement the statistical hypothesis tests in Table 1.

To summarize the Monte Carlo simulation results, we conducted visual interpretation of the model results summarized in graphs using the “marginaleffects” R package and the functions contained within it.^12^ First, we fit a logistic regression model with multiplicative interactions, as described above. We plotted how predictions and slopes change across different values of the predictors using two plotting functions: one which plots conditional adjusted predictions and another which computes conditional marginal effects. Using the first function, we generated conditional adjusted predictions of the probability of an outbreak of 50+ for the predictor variables and ranges we wanted to investigate while fixing all of the other predictors to their mean values and plotted these predictions. Similarly, using the second function, we estimated the partial derivative of outbreak of 50+ with respect to test frequency for various predictor variables and ranges we wanted to investigate while fixing all of the other predictors to their mean values, and plotted these partial derivatives.

For graphical interpretation of two-way interactions, we generated plots of the probability of an outbreak at various combinations of test frequencies from zero to four tests per week, with the key epidemiological parameter of interest held constant at informative cut points, illustrating how the slope for test frequency varied according to the second variable.

The logistic regression model included three-way interactions between test frequency, asymptomatic share, and the symptomatic self-isolation rate as well as between test frequency, pre-symptomatic share of infectious time, and the symptomatic self-isolation rate. We included these three-way interactions to characterize relationships where the two-way interaction between test frequency and a second variable was anticipated to vary across levels of a third variable. For graphical interpretation of three-way interactions, we generated faceted plots of the probability of an outbreak at various combinations of test frequency and a second epidemiological parameter, with each facet of the figure holding the third parameter constant at a different informative cut point. In so doing, we were able to illustrate how the slope for test frequency varied according to both the second and third parameters.

## Results

### Test Frequency-Transmissibility (Tau) Interaction

Figure 3.2 illustrates the interaction between test frequency and pathogen transmissibility, which is governed by tau, the per-time step probability of infection transmission across a given pair of susceptible and infectious individuals who come into contact. The plot illustrates that the relative risk reductions observed were inversely correlated with transmissibility. The absolute risk reductions observed from testing were fairly consistent across the levels tau=0.01, 0.0125, and 0.015, but shrank at tau values lower than 0.01, most likely due to floor effects. Absolute risk reductions were largest when transmissibility was at the median and second-highest cut points in the set of five. Table 2 characterizes the observed relationship between tau values and R0.

**Figure 3.2:**
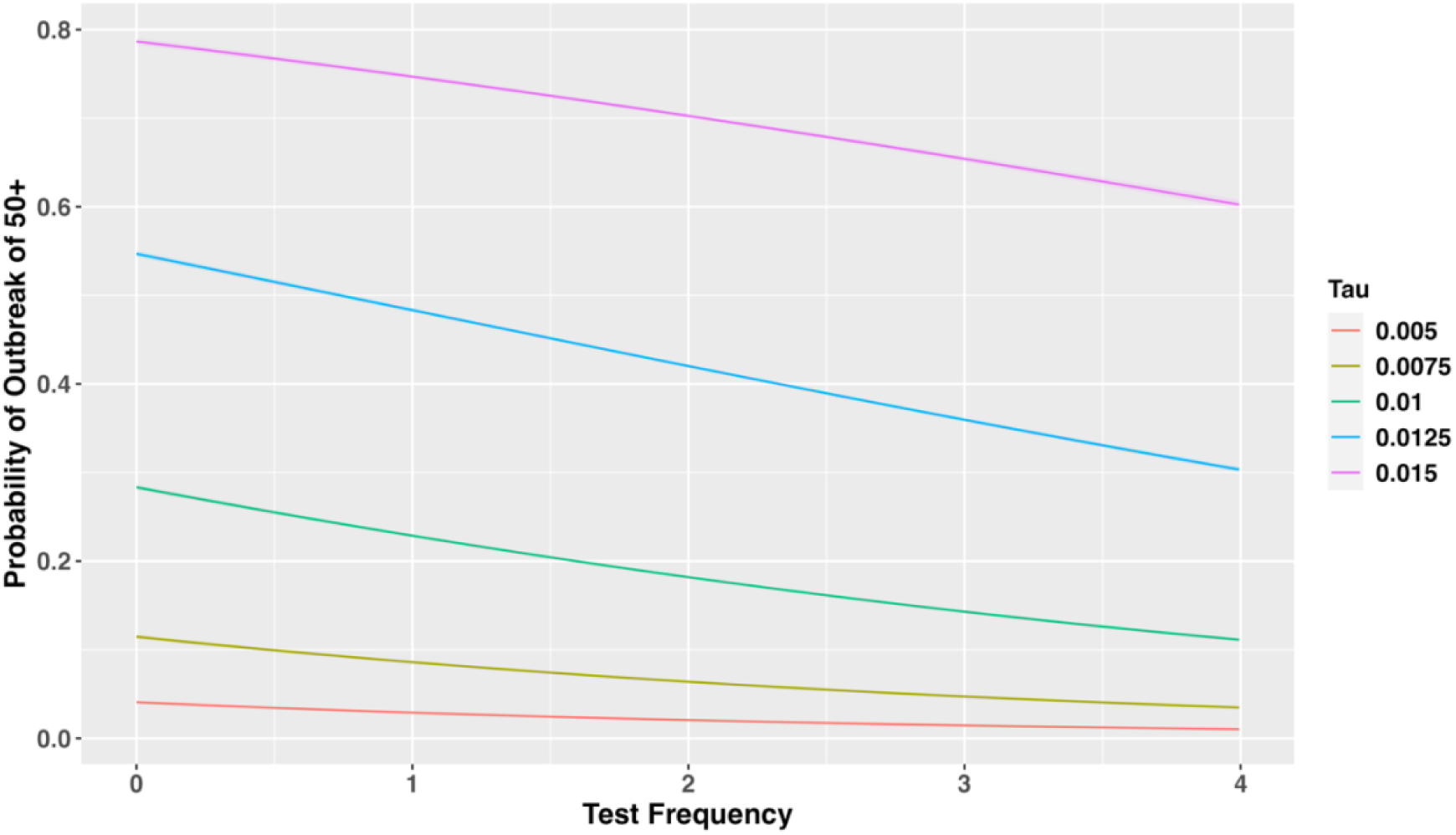
Test Frequency, Transmissibility, and Probability of an Outbreak of 50 or More Infections.

### Test Frequency, Asymptomatic Share, and Symptomatic Self-Isolation Rate

Figures 3.3, 3.4, 3.5, and 3.6 depict three-way interaction between test frequency, the symptomatic self-isolation rate, and the asymptomatic share of infections. Figure 3.7 depicts two-way interaction between test frequency and the symptomatic self-isolation rate. Two important inferences can be draw from these plots.

**Figure 3.3:**
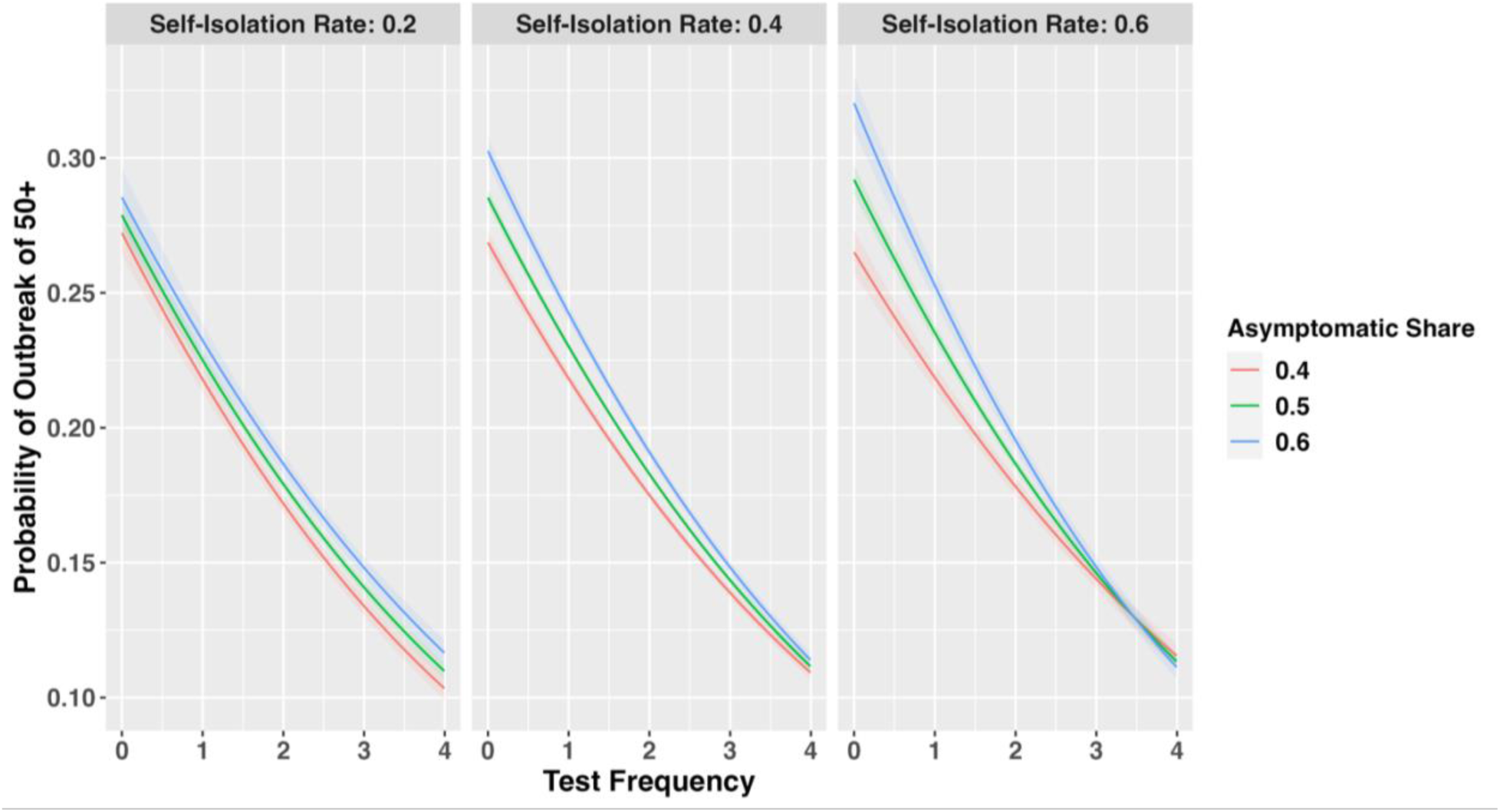
Test Frequency, Asymptomatic Share, Symptomatic Self-Isolation Rate, and Probability of an Outbreak of 50 or More Infections.

**Figure 3.4:**
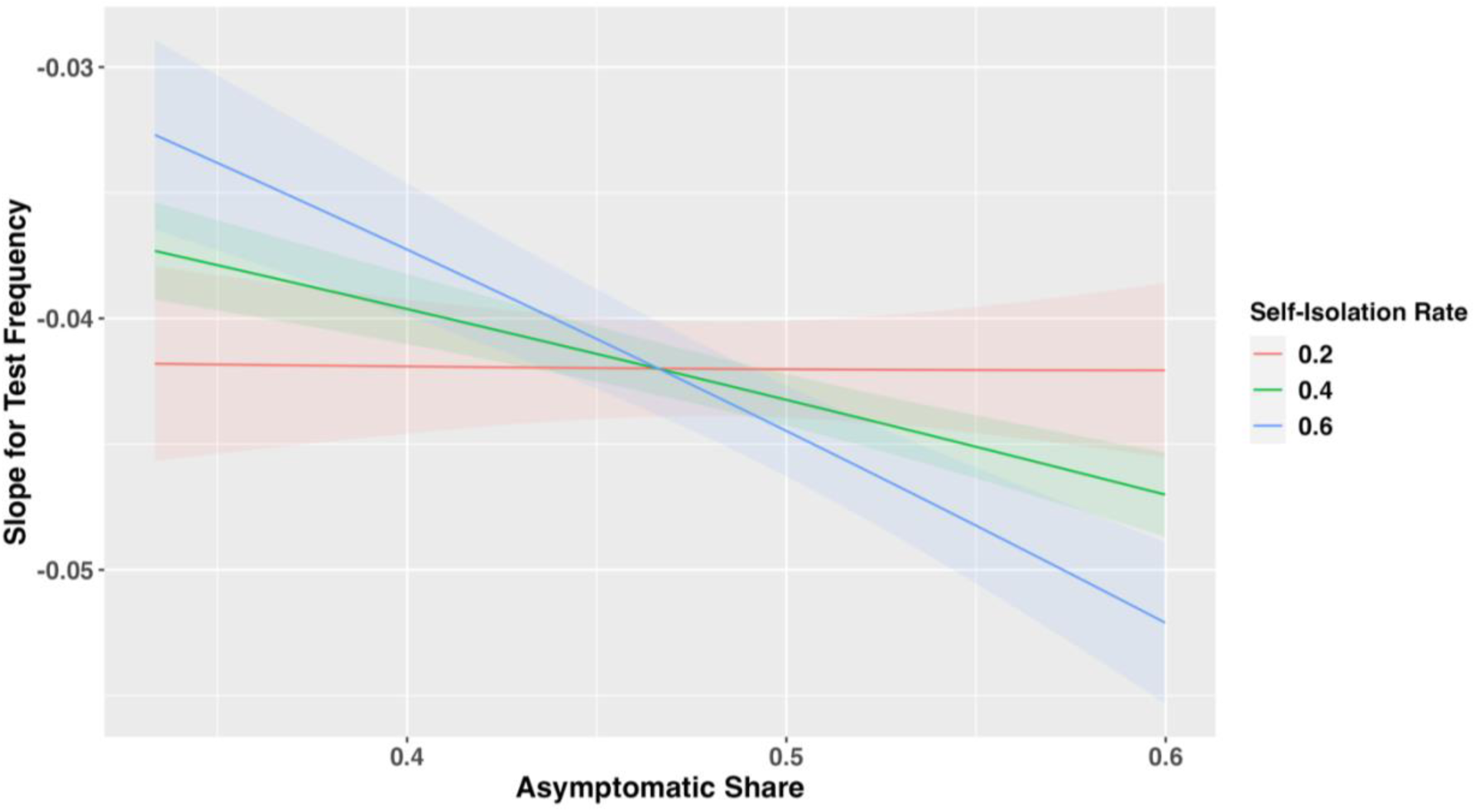
Test Frequency, Asymptomatic Share, Symptomatic Self-Isolation Rate, and Partial Derivative of the Probability of an Outbreak of 50 or More Infections.

**Figure 3.5:**
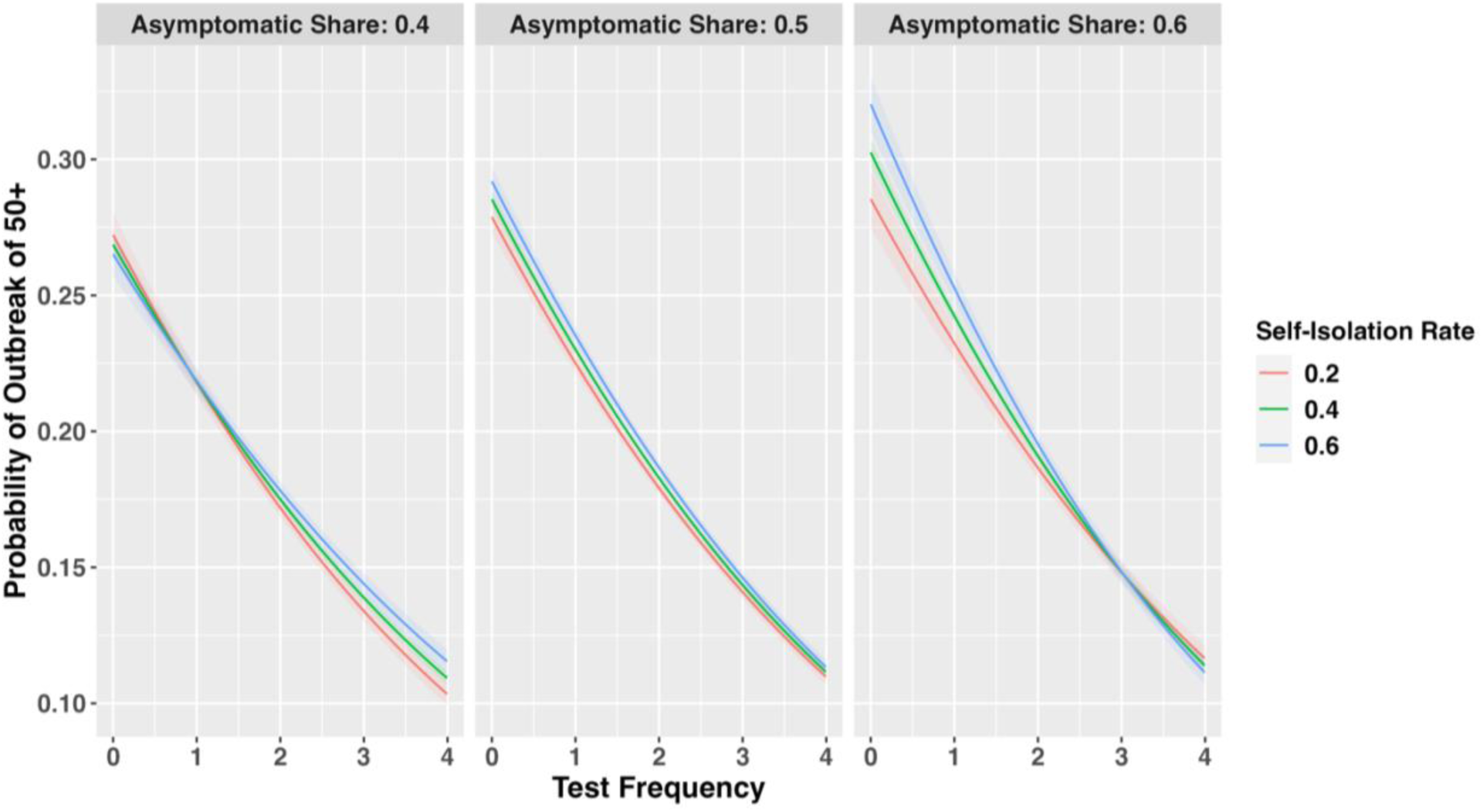
Test Frequency, Symptomatic Self-Isolation Rate, Asymptomatic Share, and Probability of An Outbreak of 50 or More Infections.

**Figure 3.6:**
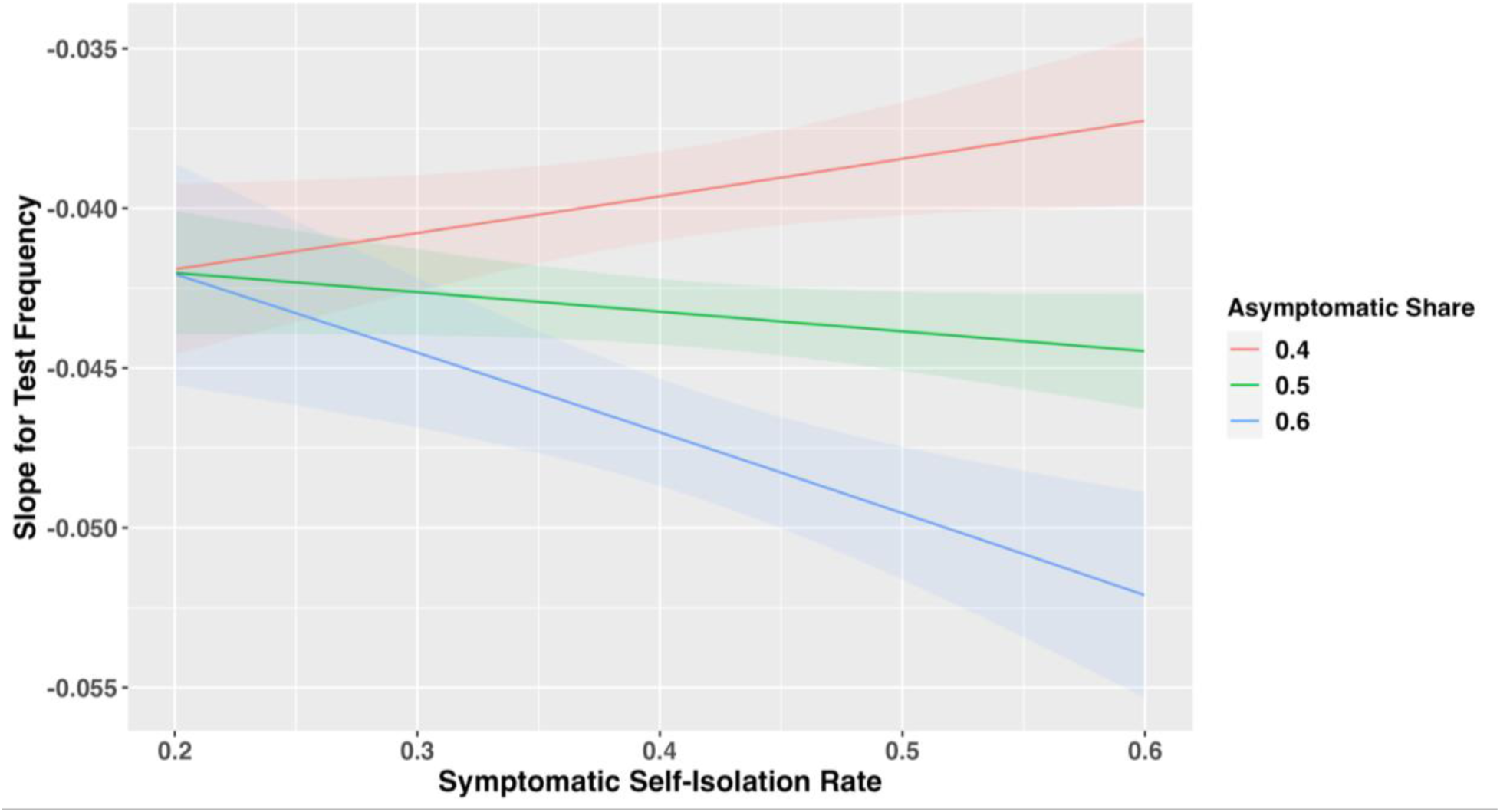
Test Frequency, Symptomatic Self-Isolation Rate, Asymptomatic Share, and Partial Derivative of Probability of an Outbreak of 50 or More Infections.

**Figure 3.7:**
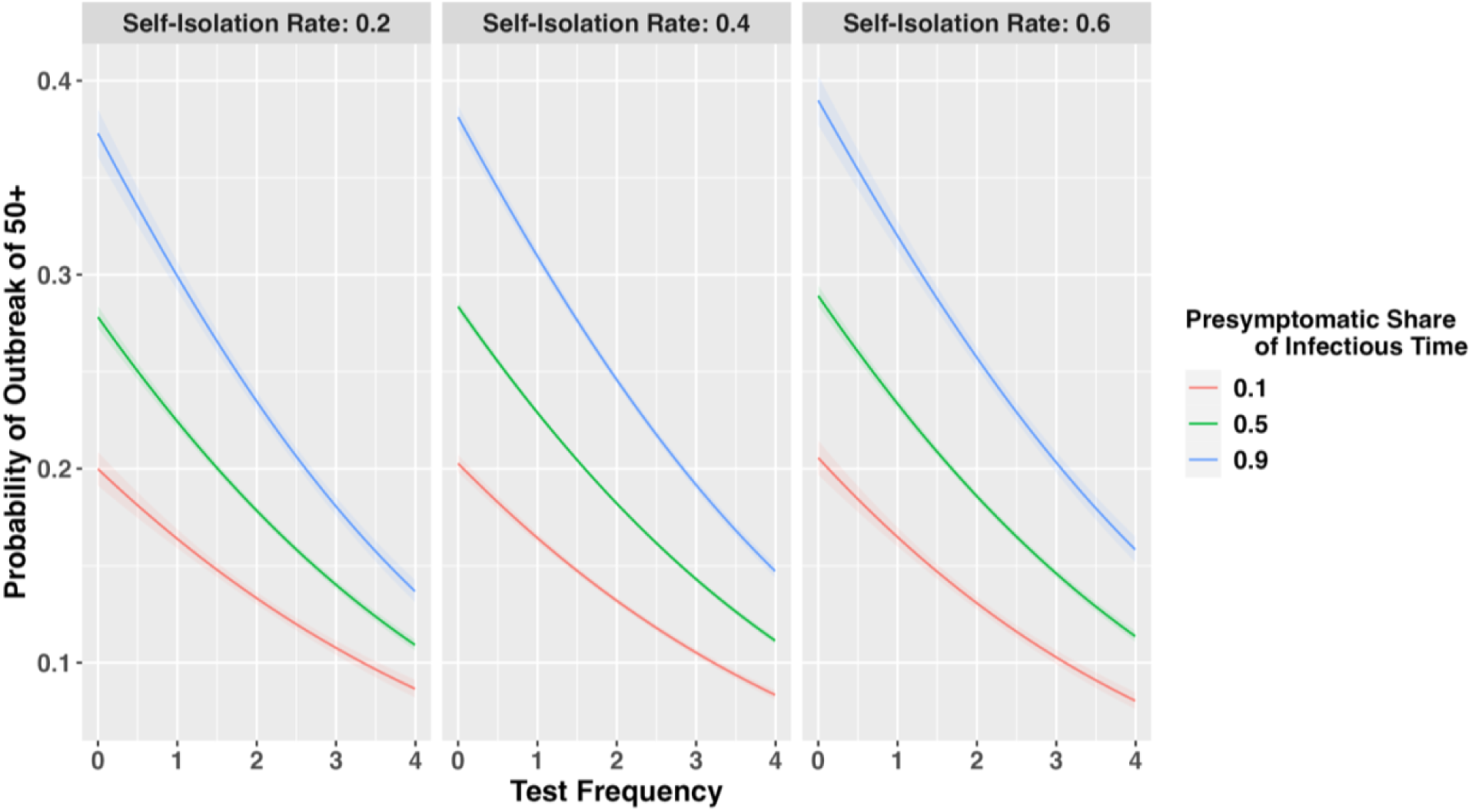
Test Frequency, Presymptomatic Share of Infectious Time, Self-Isolation Rate, and Probability of an Outbreak of 50 or More Infections.

First, Figure 3.3 and Figure 3.4 show that the effect of test frequency was magnified at higher asymptomatic shares when the self-isolation rate was high, but minimally when the self-isolation rate is low. The increased asymptomatic share increased the effect of test-induced isolation because it reduced the number of symptomatic people who might self-isolate instead, but this effect was only visible when the per-day self-isolation rate of symptomatic individuals was high enough to be a meaningful competing risk for test-induced isolation. The visual interpretation is reinforced by a significant finding in the hypothesis test for three-way interaction between these variables in Table 3.1 (p=0.001).

**Table 3.1:** Logistic Regression With Three-Way Interactions, Predicting Outbreaks of 50 or More Infections.

|  |  |  |  |  |  |
| --- | --- | --- | --- | --- | --- |
|  | Odds Ratios | Lower Bound 95% CI | Upper Bound 95% CI | Z-Score | P-Value |
| Frequency | 0.95 | 0.94 | 0.97 | -6.26 | <0.001 |
| Tau Per 100 | 1.56 | 1.55 | 1.57 | 159.91 | <0.001 |
| Symptomatic Self-Isolation Rate | 0.89 | 0.82 | 0.97 | -2.65 | 0.008 |
| Asymptomatic Share | 0.98 | 0.92 | 1.05 | -0.51 | 0.609 |
| Presymptomatic Share of Infectious Time | 1.11 | 1.09 | 1.14 | 9.99 | <0.001 |
| Frequency X Tau Per 100 | 1.01 | 1.01 | 1.02 | 10.19 | <0.001 |
| Frequency X Symptomatic Self-Isolation Rate | 1.05 | 1.01 | 1.09 | 2.64 | 0.008 |
| Frequency X Asymptomatic Share | 1.03 | 1 | 1.06 | 2.24 | 0.025 |
| Frequency X Presymptomatic Share of Infectious Time | 0.99 | 0.98 | 0.99 | -3.22 | 0.001 |
| Symptomatic Self-Isolation Rate X Asymptomatic Share | 1.29 | 1.1 | 1.51 | 3.11 | 0.002 |
| Symptomatic Self-Isolation Rate X Presymptomatic Share of Infectious Time | 1.01 | 0.96 | 1.06 | 0.44 | 0.661 |
| Frequency X Asymptomatic Share X Symptomatic Self-Isolation Rate | 0.89 | 0.83 | 0.95 | -3.47 | 0.001 |
| Frequency X Presymptomatic Share of Infectious Time X Symptomatic Self-Isolation Rate | 1.02 | 1 | 1.04 | 1.55 | 0.122 |
| Intercept | 0.55 | 0.53 | 0.58 | -31.61 | <0.001 |
| *Note: for ease of coefficient interpretation, the Tau parameter (the per time-step probability of transmission across an S-I edge) was scaled to Tau Per 100 (the number of transmission events per 100 S-I edges per time-step). |  |  |  |  |  |

**Table 3.2:** Estimated R0 Values Corresponding to Different Values of Tau.

| **Table 3.2: Estimated R0 Values Corresponding to Different Values of Tau** |  |  |  |  |  |
| --- | --- | --- | --- | --- | --- |
| **Tau:**<br>Per time-step<br>probability of<br>transmission<br>across an S-I edge | **Tau Per 100:**<br>Number of<br>transmission<br>events per 100 S-I<br>edges per time-step | **Mean R0**<br>**Week 1** | **Median R0**<br>**Week 1** | **Mean R0**<br>**Week 2** | **Median R0**<br>**Week 2** |
| **0.005** | **0.5** | 1.51 | 0.58 | 1.47 | 0.84 |
| **0.0075** | **0.75** | 1.82 | 1.4 | 1.74 | 1.39 |
| **0.01** | **1** | 2.14 | 1.4 | 2.02 | 1.4 |
| **0.0125** | **1.25** | 2.46 | 1.58 | 2.27 | 1.4 |
| **0.015** | **1.5** | 2.8 | 2.23 | 2.57 | 2.12 |
| Note: simulations were conducted with no routine testing that systematically varied only tau while fixing the asymptomatic share at 0.5, the pre-symptomatic share of infectious time at 0.5, and the symptomatic self-isolation rate per day at 0.4 |  |  |  |  |  |

Second, Figure 3.5 and Figure 3.6 illustrate that the direction of how the symptomatic self-isolation rate moderated the effect of increased test frequency depended on the asymptomatic share. The effect of increased test frequency was attenuated at higher self-isolation rates when the asymptomatic share was lower presumably because when self-isolation rates were high and most cases were symptomatic, self-isolation represented a substantial competing risk to test-induced isolation. In contrast, the effect of increased test frequency was magnified at higher self-isolation rates when the asymptomatic share was higher. This may have occurred because when the asymptomatic share was high and self-isolation rate was high, there were more people who counterfactually would have self-isolated had they shown symptoms but end up being isolated as a result of testing instead, compared to a counterfactual scenario where the self-isolation rate was lower, but the asymptomatic share was still high.

### Test Frequency, Pre-symptomatic Share of Infectious Time, and Symptomatic Self-Isolation Rate

Figure 3.7 and Figure 3.8 depict a hypothesized three-way interaction between test frequency, the pre-symptomatic share of infectious time, and the symptomatic self-isolation rate. Figure 3.7 and Figure 3.8 illustrate that as the pre-symptomatic share of infectious time increases, the effect of test frequency on the probability of an outbreak is strongly magnified. This is expected because as fewer infectees show symptoms, fewer will self-isolate, leaving more infectees available for test-induced isolation. Figure 3.8 illustrates that while the symptomatic self-isolation rate slightly moderates the effect of test frequency when very little infectious time is pre-symptomatic (most time is symptomatic), the symptomatic self-isolation rate does not moderate the effect of test frequency when most infectious time is pre-symptomatic, most likely because people need to show symptoms in order for the symptomatic self-isolation rate to exert an effect. The absence of a practically significant three-way interaction between these variables is reflected in the absence of a statistically significant finding of three-way interaction in the hypothesis test shown in Table 3.1 (p=0.12). From a practical perspective, these findings indicate that the pre-symptomatic share of infectious time magnifies the effect of test frequency largely independently of the symptomatic self-isolation rate.

**Figure 3.8:**
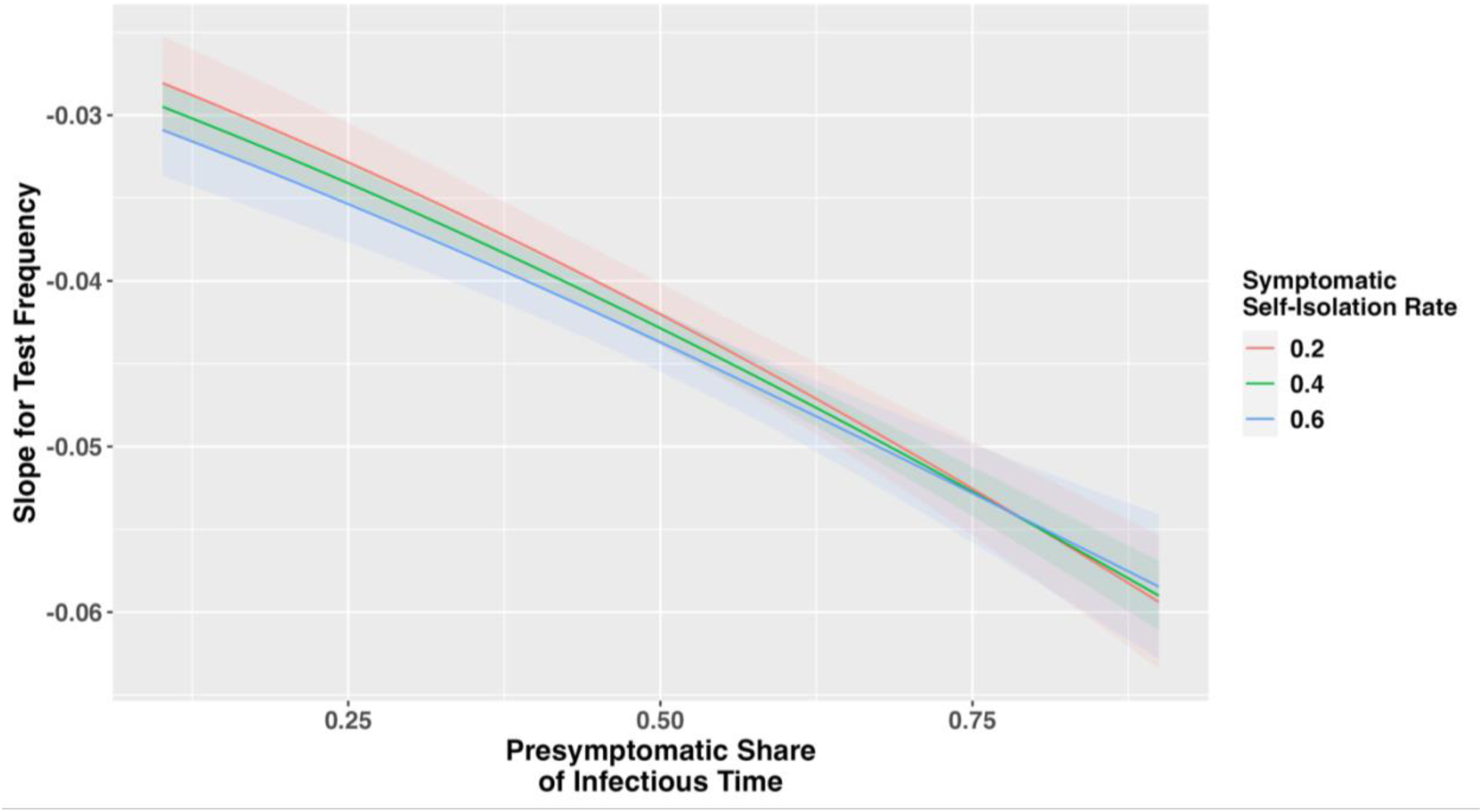
Test Frequency, Presymptomatic Share of Infectious Time, Symptomatic Self-Isolation Rate, and Partial Derivative of Probability of an Outbreak of 50 or More Infections.

**Figure 3.9:**
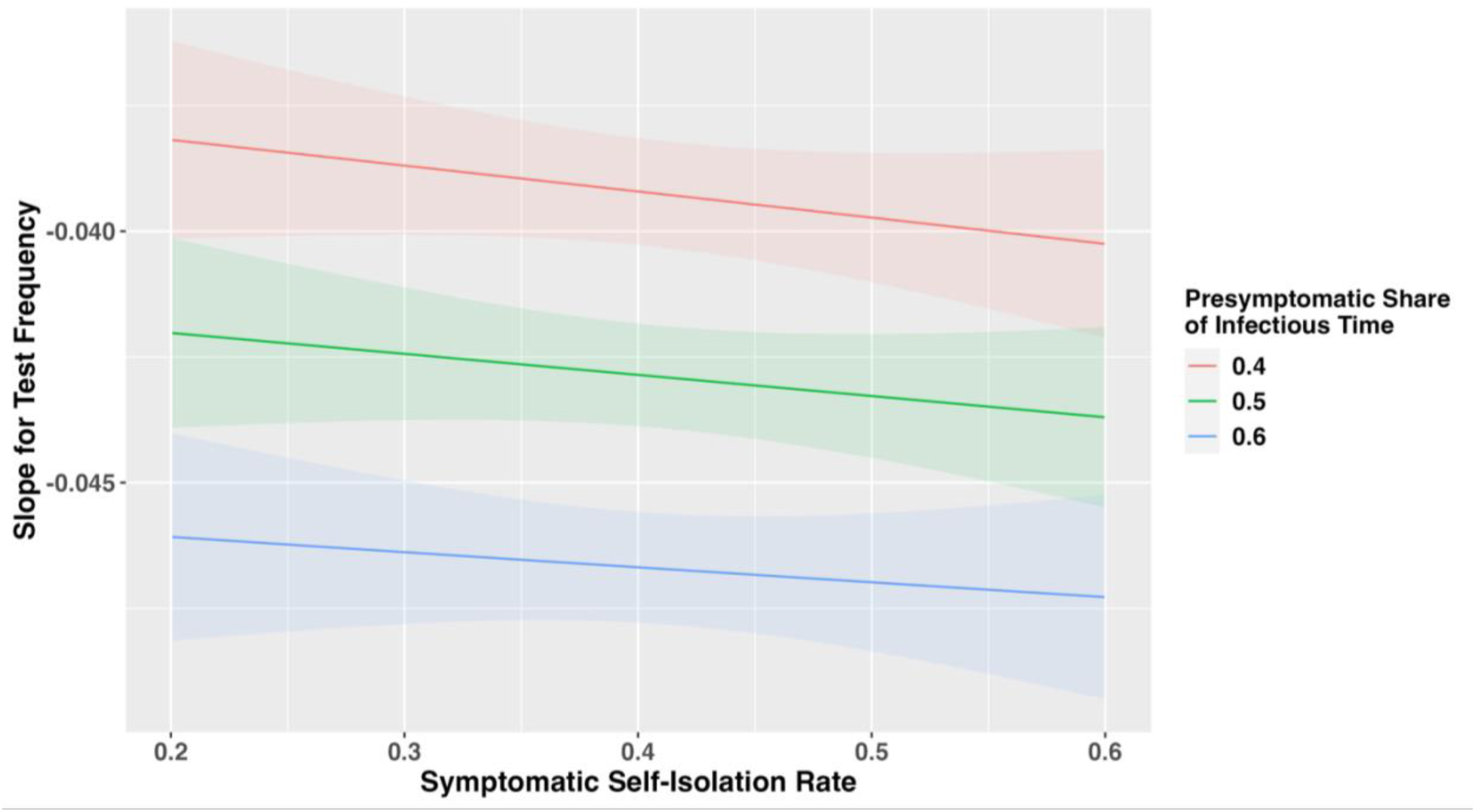
Test Frequency, Presymptomatic Share of Infectious Time, Symptomatic Self-Isolation Rate, and Partial Derivative of Probability of an Outbreak of 50 or More Infections.

## Discussion

In this study we modeled how the epidemiological characteristics of PPPs moderate the efficacy of a routine testing and isolation intervention in preventing larger outbreaks after an LAI. We used a discrete-time stochastic network infectious disease model to run 625,000 epidemic simulations encompassing 625 unique combinations of test frequency, pathogen transmissibility, the self-isolation rate for symptomatic cases, the percentage of cases that are asymptomatic, and the percentage of infectious time that is spent in the pre-symptomatic state among those who show symptoms. To summarize our Monte Carlo simulations, we combined visual analysis with logistic regression for formal hypothesis testing, focusing on the interaction terms that capture the moderating effect of epidemiological parameters on the impact of test frequency.

The first key finding is that the relative risk reductions of PPP escape observed from increased test frequency were inversely correlated with pathogen transmissibility. This is consistent with the notion that that as pathogen transmissibility rises, the relative risk reductions that can be achieved with increased test frequency decline due to the inherent difficulty of containing a highly transmissible pathogen. However, this should not be misunderstood to mean that testing for the most transmissible pathogens is not worth conducting given the difficulty of containing them after an initial infection. In fact, the observed absolute risk reductions achieved from testing are fairly consistent across the middle to highest levels of pathogen transmissibility modeled. Moreover, the level of societal disruption caused by a pathogen escape would likely rise along with increases in pathogen transmissibility, in part because an epidemic caused by a pathogen with higher transmissibility will have a faster, higher peak. The steepness and peak of the initial surge in cases during an epidemic is an important determinant of the ability to maintain critical services such as the health care system.^13^ Moreover, even if the testing and isolation intervention modeled proved inadequate for containment, early detection of a pathogen remains crucial for ensuring a rapid public health response.^14^

The second key finding is that the effect of test frequency was magnified at higher asymptomatic shares when the symptomatic self-isolation rate was high, but minimally when the self-isolation rate was low. The increased asymptomatic share increased the effect of test-induced isolation because it reduced the number of symptomatic people who might self-isolate instead, but this effect was only visible when the per-day self-isolation rate of symptomatic individuals was high enough to be a meaningful competing risk for test-induced isolation. The third key finding is that the direction of how the symptomatic self-isolation rate moderated the effect of increased test frequency depended on the asymptomatic share. The effect of increased test frequency was attenuated at higher self-isolation rates when the asymptomatic share was lower and was magnified at higher self-isolation rates when the asymptomatic share was higher. The fourth key finding is that as the pre-symptomatic share of infectious time increased, the effect of test frequency on the probability of an outbreak was strongly magnified. This was expected because as fewer infectees showed symptoms, fewer self-isolated, leaving more infectees available for test-induced isolation.

From a policy perspective, these other key findings tell us several things. First, a routine testing and isolation intervention is likely to be especially valuable in reducing the risk of a catastrophic lab escape of potential pandemic pathogens that exhibit a high share of asymptomatic cases. A number of known pandemic pathogens exhibit asymptomatic transmission, including SARS-COV-2 and influenza, and could grow to include future pandemic pathogens whose transmission patterns in humans have yet to emerge or be characterized.^15^ Second, such an intervention is also likely to be especially valuable in reducing the risk of a catastrophic lab escape of potential pandemic pathogens for which a significant share of infectees’ infectious time is spent pre-symptomatically, among those infectees who will develop symptoms. This category also includes SARS-COV-2, and may come to include others as well.^15^

The US government policy frameworks that currently govern dual use research of concern with high-consequence pathogens are designed to subject all potentially high-risk NIH-funded research involving pathogens with enhanced pandemic potential (PEPPs) to departmental-level risk-benefit review by the U.S. Department of Health and Human Services.^16^ ^17^ ^18^ ^19^ However, under these policies, the criteria that define which pathogens with enhanced pandemic potential for which wet-lab research is automatically subject to departmental-level review do not explicitly take into account the degree of a pathogen’s transmission that is asymptomatic or pre-symptomatic. This study’s findings indicate that the degree of asymptomatic or pre-symptomatic transmission is an important determinant of the feasibility of accidental escape prevention, even in the context of a vigorous lab worker health surveillance regime. This factor should be incorporated into future risk assessments to determine whether routine testing of lab workers would be necessary and appropriate in a particular context. Moreover, future risk assessments should focus not only on a pathogen’s current characteristics, but also the new characteristics that the research activities are intended to or are reasonably anticipated to provide it with.^19^ Insofar as pathogen modification might be reasonably anticipated to enhance the degree of asymptomatic or pre-symptomatic transmission, this should also be taken account of in such risk assessments.

The self-isolation rate of symptomatic infected lab workers is potentially an important target of policy intervention. Lab workers’ symptomatic self-isolation rate is likely a function of a variety of factors. While the published literature on the determinants of self-isolation among lab workers after lab-acquired infections is sparse, likely due to the rarity of these events, there is a related literature on the factors that influence compliance with quarantine policies during infectious disease outbreaks. Two key factors that influence quarantine compliance are the perceived risks of disease transmission and severity as well as the perceived benefit of compliance.^20^ These factors could conceivably be influenced by lab worker biosafety training programs and thus influence the self-isolation rate. Another factor known to influence quarantine compliance is the practical difficulty of compliance in the face of competing considerations such as foregone wages, family care obligations, securing supplies, and psychological stress from isolation.^20^ ^21^ Previous research found a mandatory paid sick leave policy increased compliance with self-isolation and quarantine measures to control the COVID-19 epidemic,^22^ suggesting that similar policies to provide economic and social support to self-isolating lab workers could increase the symptomatic self-isolation rate. Similarly, social norms, cultural values, and legal requirements have also been found to influence quarantine compliance during epidemics.^20^ Policymakers might aim to boost lab workers’ symptomatic self-isolation rate by targeting these factors through education, persuasion, and regulation. Another factor that may influence the symptomatic self-isolation rate is the perceived probability that symptoms could be caused by a lab-acquired infection with a potential pandemic pathogen rather than something else, given that the lab workers will encounter other pathogens and sources of illness in their daily lives. Here, too, policies to educate lab workers on the risks of accidental lab-acquired infections that threaten not just themselves, but society at large might shift lab workers more towards adopting a precautionary presumption towards self-isolation, even in the absence of confirmatory testing.

## Conclusion

This epidemiological modeling study finds that while routine testing of lab workers in high biosafety research facilities coupled with isolation of infected lab workers can reduce the risk of a catastrophic lab escape of a potential pandemic pathogen, the efficacy of such an intervention will be mediated by a pathogen’s epidemiological characteristics. Key epidemiological characteristics include the pathogen’s transmissibility, the symptomatic self-isolation rate, the asymptomatic share of total infections, and the pre-symptomatic share of infectious time among those who show symptoms. This modeling study finds that the relative risk reductions observed from increased test frequency are inversely correlated with transmissibility. It also finds that the effect of test frequency is magnified at higher asymptomatic shares when the symptomatic self-isolation rate is high, but hardly at all when the self-isolation rate is low. Moreover, the study finds that the direction of how the symptomatic self-isolation rate moderates the effect of increased test frequency depends on the asymptomatic share, attenuating it when the asymptomatic share is low and magnifying the effect when the asymptomatic share is high. Additionally, the study finds that as the pre-symptomatic share of infectious time increases, the effect of test frequency on the probability of an outbreak is strongly magnified. These findings indicate that a routine testing and isolation intervention is likely to be especially valuable in reducing the risk of a catastrophic lab escape of potential pandemic pathogens that exhibit higher shares of asymptomatic and pre-symptomatic transmission, and that lab workers’ symptomatic self-isolation rate is likely to be an important target of policy intervention.

## Data Availability

All data produced in the present study are available upon reasonable request to the authors

